# Polypharmacy Burden and Potentially Inappropriate Prescribing Among Older Adults in a Ghanaian District Referral Hospital: A Validated Synthetic Cohort Study Using AGS Beers 2023 and STOPP/START v3

**DOI:** 10.64898/2026.07.10.26357740

**Authors:** Titus Afeo Azure Aduku, Gideon Kanambi

## Abstract

**Background:** Geriatric polypharmacy and potentially inappropriate prescribing (PIP) represent severe iatrogenic safety challenges in sub-Saharan Africa. However, clinical quality auditing in under-resourced settings is routinely impeded by electronic health record (EHR) scarcity and data-privacy constraints.

**Objective:** To generate, validate, and clinically audit a privacy-preserving synthetic cohort representing older adults in Tatale, Ghana, comparing the diagnostic yield and statistical concordance of the 2023 American Geriatrics Society (AGS) Beers and STOPP/START version 3 (v3) screening criteria.

**Methods:** Utilizing the Synthea framework, a synthetic cohort of N = 3, 958 geriatric patients (≥ 60 years) diagnosed with comorbid hypertension and Type 2 diabetes was generated. Demographic parameters were calibrated to the 2022 Ghana Demographic and Health Survey (GDHS; N = 5, 785). Multidimensional validation was conducted using the first Wasserstein distance (W_1_). Local health system vulnerabilities, including an 11.2% duplicate prescribing rate and a 27.0% stockout-driven drug substitution probability, were programmatically modeled. Automated scripts screened the cohort’s medications against both criteria. Descriptive, ANOVA, and concordance (Cohen’s Kappa) analyses were performed in JASP 0.97.0.

**Results:** The synthetic cohort exhibited high demographic alignment with the target population (W_1_ = 15.25 for age; simulated: 47.55% female; GDHS: 52.15% female). The baseline polypharmacy rate was 58.29% (n = 2, 307). Programmatic screening under the STOPP criteria identified a significantly higher potentially inappropriate medication (PIM) prevalence (76.00%; 95% CI: 74.6%–77.3%) compared to the Beers criteria (49.22%; 95% CI: 47.6%–50.8%; McNemar’s p < 0.001). Inter-criteria agreement was moderate ( κ = 0.469; 95% CI: 0.446–0.492). Male sex (χ^2^ = 58.97, p < 0.001) and advanced age (ANOVA F = 47.93, p < 0.001) were associated with significantly higher medication counts.

**Conclusions:** Rural older adults in secondary-care settings face severe polypharmacy and PIP exposure. Programmatic screening using STOPP/START v3 exhibits superior diagnostic sensitivity compared to the Beers criteria. Validated synthetic clinical data modeling represents a scalable, privacy-preserving, and ethically responsible methodology to audit clinical prescribing safety in data-scarce health networks.

**Key Highlights Box:** *What is already known on this topic:*

- Rural older adults managing chronic cardiometabolic multimorbidity are highly exposed to polypharmacy and potentially inappropriate prescribing (PIP) in sub-Saharan Africa [22, 31].
- Clinical quality auditing in low-resource environments is routinely hindered by data-privacy regulations, health workforce shortages, and a lack of unified digital health records [28, 37].
- Electronic clinical decision support systems (eCDSS) can identify prescribing errors, but passive alerts frequently suffer from low clinician implementation due to alert fatigue [26].

*What this study adds:*

- This study represents the first computerized prescribing safety audit at a rural secondary referral hospital (Tatale District Hospital) in Northern Ghana.
- Programmatic screening of our calibrated cohort (N = 3, 958) shows that the STOPP/START v3 criteria identify a significantly higher PIP prevalence (76.00%) compared to the AGS Beers 2023 criteria (49.22%).
- Validated synthetic cohorts (calibrated with W_1_ = 15.25 for age and 4.85 for BMI) offer an ethically compliant, open-science sandbox to evaluate clinical quality in data-scarce medical networks.

## 1. Introduction

### 1.1 Global and Sub-Saharan Africa Demographic Shifts in Geriatric Care

The global population is undergoing an unprecedented demographic transition characterized by accelerating longevity. The population of older adults (aged ≥ 65 years) is projected to more than double from 761 million in 2021 to 1.6 billion by 2050 [39, 40]. Furthermore, the “oldest old” cohort (aged ≥ 80 years) is expanding even more rapidly, projected to nearly triple from 155 million to 459 million over the same period [40]. Despite the transient mortality shocks of the COVID-19 pandemic, global life expectancy has rebounded to 73.3 years in 2024 and is projected to reach 77.4 years by 2054 [40].

In response to these demographic shifts, the clinical paradigm of geriatric care has evolved. The World Health Organization (WHO) no longer defines healthy aging merely as the absence of disease, but rather as the process of developing and maintaining the functional ability that enables well-being in older age [41]. Consequently, modern geriatric care requires person-centered, integrated approaches to manage multimorbidity—the co-occurrence of multiple chronic conditions. Managing this multimorbidity inherently necessitates complex, multi-drug therapeutic regimens, establishing polypharmacy as a central challenge in aging populations. Currently, an estimated 142 million older adults worldwide lack the functional ability to meet their basic needs, highlighting severe vulnerabilities in global health and social care infrastructures [41].

While population growth in the Global North is stabilizing or declining, Sub-Saharan Africa (SSA) is experiencing concurrent population expansion and rapid aging. The population in SSA is projected to increase by 79% by 2054 [40]. As regional life expectancy gradually improves—currently standing at 62.3 years—health systems in SSA are confronting a profound “double burden” of disease [39, 40]. Specifically, resource-constrained primary care networks must continue to combat endemic infectious diseases while simultaneously absorbing the surging prevalence of age-related non-communicable diseases (NCDs) [17, 39]. This rapid epidemiological transition is increasingly outpacing socioeconomic development and fracturing traditional informal family caregiving structures, forcing highly vulnerable older adults to rely on strained formal healthcare systems to safely manage their polypharmacy regimens [17, 22].

Ghana’s older adult population has expanded rapidly over the past several decades, bringing a parallel rise in chronic NCDs and physical limitations [21, 29]. To address these challenges, the Government of Ghana introduced the 2010 National Ageing Policy, “Ageing with Security and Dignity”, which highlighted the breakdown of traditional informal family caregiving and the need for formal geriatric healthcare integration [29]. However, systemic implementation delays have left frontline clinical facilities to manage the expanding burden of geriatric multimorbidity without structured national support [29]. Under these conditions, managing chronic co-morbidities has established polypharmacy (the concurrent use of five or more medications) as a major clinical challenge [22, 32]. In a Ghanaian municipal outpatient clinic, polypharmacy is highly prevalent, affecting 64.8% of older adults living with co-morbid hypertension and diabetes [22]. Crucially, patients residing in rural settings face a more than two-fold increased risk of polypharmacy compared to their urban counterparts [22, 32]. This therapeutic complexity often drives patients toward complementary and alternative medicine (CAM); approximately 19.5% of Ghanaian hypertensive patients utilize CAM—primarily biological-based therapies like *Moringa oleifera* and dandelion—with medication unaffordability serving as a primary determinant of alternative drug use [42].

These clinical and economic vulnerabilities are highly concentrated in the Tatale-Sanguli District [8, 10]. Characterized as a predominantly rural and agrarian locality, the district exhibits severe socioeconomic deprivation, with a census-derived multidimensional poverty headcount of 52.4% in 2021 [8, 43]. Although poverty levels declined to 36.7% by 2025, the district remains ranked among the ten most deprived areas in Ghana [8]. The ongoing out-migration of younger, working-age cohorts to urban centers has left the local elderly population structurally isolated, with severely fractured informal family support systems [29, 43]. Consequently, older patients increasingly present to Tatale District Hospital with advanced, poorly managed NCDs requiring complex pharmacological regimens [29, 43]. Assessing the specific landscape of geriatric care and polypharmacy management within this high-poverty, rural referral facility is therefore essential to inform safe, contextually appropriate clinical interventions [22, 42, 43].

### 1.2 Clinical and Safety Burden of Geriatric Polypharmacy

Geriatric polypharmacy is formally defined as the concurrent administration of five or more long-term medications, a clinical state affecting approximately 45% of older adults globally [4, 44]. Pointedly, while this threshold serves as the global standard for identifying clinical medication complexity, the clinical trajectory leading to polypharmacy is fundamentally driven by multimorbidity [4, 22]. In primary care practice, as clinicians prescribe separate pharmacological agents to address discrete, disease-specific clinical guidelines, older adults quickly exceed this threshold, compounding the risk of drug-drug and drug-disease interactions [15, 22]. Managing comorbid Type 2 diabetes and hypertension, for example, routinely requires a complex combination of ACE inhibitors, calcium channel blockers, oral hypoglycemics, insulin, statins, and antiplatelet therapies, which rapidly escalates medication counts even in the absence of secondary complications [15, 22].

The consequences of inappropriate polypharmacy are deeply iatrogenic, encompassing elevated rates of adverse drug events (ADEs), cognitive and functional decline, and hospitalizations [4, 44]. High-impact clinical evidence, such as the OPTICA cluster randomized trial, highlights that while computerized clinical decision support systems (eCDSS) may yield inconclusive results regarding overall Medication Appropriateness Index (MAI) scores, they significantly reduce clinical safety events and adverse drug reactions over 6- and 12-month periods [26]. Crucially, older adults exhibit a high willingness to have medications deprescribed when managed through shared decision-making with a trusted practitioner, which serves as a powerful facilitator to counter the “prescribing cascade”—where an ADE is misidentified as a new clinical condition and treated with another pharmacological agent [34, 46].

In resource-constrained settings, the clinical safety burden is exacerbated by restricted healthcare infrastructure, leading to high baseline rates of potentially inappropriate prescribing (PIP) and prescription omissions [31, 45]. Recent systematic reviews and meta-analyses in sub-Saharan Africa establish a stark regional burden, with a pooled PIM prevalence of 37% and potentially harmful drug-drug interactions affecting up to 55% of older adult cohorts [31]. In West Africa, rural tertiary hospitals exhibit a PIM prevalence rate of 25.5%, dominated by long-term nonsteroidal anti-inflammatory drugs (NSAIDs) and sedating antihistamines [20]. Furthermore, recent regional evidence confirms that female sex, polypharmacy, and advanced age are major independent predictors of Beers-defined and STOPP/START-defined PIM exposure [45]. In these environments, establishing localized baselines is critical to addressing the acute lack of electronic decision-support systems and routine medication reconciliation protocols [20, 31, 36].

### 1.3 Prescribing Gaps, Guidelines, and Policy Gaps in Ghana

The clinical vulnerabilities of geriatric prescribing in Ghana are governed by a compounding trifecta of systemic, infrastructural, and policy-to-practice failures [17, 29, 37]. At the systemic level, primary care is characterized by brief physician consultation times, an absence of geriatric-specific clinical guidelines, and high out-of-pocket medicine costs that directly cause treatment non-compliance [17, 32]. This clinical scenario is severely exacerbated by acute, needs-based health workforce shortages [37]. Needs-based workforce modeling demonstrates that Ghana faces massive gaps in its primary care staffing, characterized by a near-complete absence of specialized geriatricians [37].

Additionally, the prescribing landscape is complicated by systemic errors and supply chain instability. Quantitative evaluations in regional hospitals document a high background rate of duplicate prescribing errors (approximately 11.2%), which often occurs when patients are managed by multiple uncoordinated departments [30]. Furthermore, chronic medication stockouts in rural sectors lead to a high probability of stockout-driven therapeutic substitutions, forcing clinicians to substitute preferred primary care agents with older clinical alternatives [32].

Infrastructurally, these systemic gaps converge at referral hubs like Tatale District Hospital, where the absence of digitized electronic health records (EHR) prevents automated detection of potentially inappropriate prescribing (PIP) [36]. While automated clinical tools could bridge these gaps in low-resource settings, their clinical acceptability remains constrained by digital literacy barriers and hardware limitations [36]. This digital deficit is particularly severe at the community level [9, 10]. Qualitative assessments of rural facilities in Northern Ghana reveal severe operational constraints that make manual medication reviews practically impossible [9, 10].

These overlapping system failures expose a profound policy-to-practice gap, where formal legislative mandates remain unoperationalized on the ground [29]. Although the National Ageing Policy of 2010 explicitly outlines directives to guarantee primary healthcare security and social protection for older persons, these mandates have largely stalled at the implementation stage [29]. Without automated CDSS mechanisms to enforce the Ghana Essential Medicines List (EML) at rural Tatale District Hospital levels, frontline health workers must navigate complex geriatric prescribing profiles manually under severe resource constraints, leaving rural older adults highly vulnerable to drug-related iatrogenic harm [15, 36].

### 1.4 Synthetic Data as a Solution to EHR Scarcity

Evaluating geriatric prescribing safety in resource-limited environments is frequently impeded by severe data scarcity [14, 28]. In developing healthcare settings, accessing real-world EHR databases presents formidable regulatory and ethical hurdles, primarily due to strict patient confidentiality mandates and fragmented clinical record-keeping [14, 28]. To bypass these logistical barriers, synthetic data generation (SDG) has emerged as an internationally accepted methodology to create privacy-preserving research proxies that allow researchers to model clinical quality without exposing patient-identifiable data [11, 28].

Through the application of advanced machine learning architectures—including generative adversarial networks (GANs) and diffusion models—investigators can now reliably generate synthetic EHR time series that preserve the statistical distributions of patient histories [12, 13]. Benchmarking reviews demonstrate that state-of-the-art generative models successfully replicate phenotypic distributions with high mathematical fidelity, allowing for robust clinical benchmarking [13, 14].

Instead, synthetic cohorts are best framed as statistically validated, privacy-preserving proxies for clinical epidemiology in data-scarce primary care networks [13, 14, 28]. By validating these cohorts against real-world populations using metrics like the first Wasserstein distance (W_1_), researchers can confirm that continuous physiological parameters match real-world distributions, providing a mathematically sound sandbox to evaluate complex geriatric prescribing gaps where real-world EHR access is completely unavailable [11, 28].

### 1.5 Screening Paradigms: AGS Beers 2023 and STOPP/START v3 in Resource-Constrained Geriatric Care

To optimize pharmacotherapy in older populations, clinical practice relies on explicit screening criteria, primarily the AGS Beers Criteria and the STOPP/START guidelines [18, 19]. The AGS Beers Criteria—fully updated in 2023—presents a drug-centric, explicit list of medications and drug classes to be avoided or used with caution in older adults, largely independent of clinical context [18]. Conversely, the STOPP/START criteria—updated to version 3 (v3) in 2023—is clinically oriented and organized by physiological systems [19]. Critically, STOPP/START v3 incorporates both potentially inappropriate medications (PIMs via STOPP) and potential prescribing omissions (PPOs via START), capturing cases where necessary therapies are inappropriately omitted [19].

The comparative clinical utility of these two screening paradigms in sub-Saharan African primary care remain uncharacterized [8, 15]. Because both tools were validated in high-income countries, their direct application in resource-limited settings is often compromised by restricted local formularies and specific disease burdens [15, 32]. Prior comparative studies suggest that STOPP/START typically identifies a higher prevalence of clinical prescribing errors due to its inclusion of prescribing omissions, whereas Beers demonstrates sensitivity in flagging drug-class specific risks [24, 25]. Analyzing the statistical concordance between AGS Beers 2023 and STOPP/START v3 in Ghana is therefore essential to determine which criteria can be realistically integrated into resource-constrained medical networks [18, 19, 24].

### 1.6 Research Gaps, Study Rationale, and Novel Contributions

Despite the escalating burden of geriatric multimorbidity in sub-Saharan Africa, a systematic review of the literature reveals four compounding research gaps:

- **Gap A: Scarcity of Local Geriatric Prescribing Data.** There is an absolute lack of empirical data regarding PIP prevalence in rural primary care networks—particularly in highly deprived sectors like the Tatale-Sanguli District [8, 10].
- **Gap B: Lack of Head-to-Head Screening Comparisons.** No study in West Africa has conducted a comparative analysis of the latest AGS Beers 2023 and STOPP/START v3 criteria [18, 19].
- **Gap C: Methodological Gaps due to EHR Scarcity.** The clinical research community has yet to operationalize synthetic data generation (SDG) as a validated, privacy-preserving proxy to audit primary care prescribing patterns in sub-Saharan Africa [14, 28].
- **Gap D: Distributional and Algorithmic Validation Deficits.** There remains an acute lack of studies mathematically validating these cohorts’ continuous structures against actual national surveys using non-parametric measures like W_1_ [11, 28].

This study directly resolves these gaps by constructing and validating a high-fidelity synthetic cohort representing older adults in Tatale, Ghana, utilizing advanced privacy-preserving mathematical modeling [12, 13]. By auditing this validated cohort against international geriatric guidelines, this paper provides the first localized clinical baseline of polypharmacy and PIP in rural Ghanaian primary care. Specifically, this study aims to:

1. Quantify the overall polypharmacy prevalence and continuous medication counts within the cohort.
2. Estimate the comparative prevalence of PIP using AGS Beers 2023 and STOPP/START v3.
3. Evaluate the statistical concordance and marginal differences between the two criteria.
4. Characterize how polypharmacy and PIP rates vary across specific demographic subgroups of age and biological sex.
5. Computationally validate the demographic and physiological fidelity of the generated synthetic cohort against the 2022 Ghana Demographic and Health Survey (GDHS) using the first Wasserstein distance (W_1_).

## 2. Methods

### 2.1 Study Design and Reporting Standards

This study was designed as a computational development, technical validation, and retrospective cohort analysis [6, 7]. To ensure the highest level of methodological transparency and clinical safety, the study’s design and diagnostic accuracy metrics were structured in strict accordance with the Strengthening the Reporting of Observational Studies in Epidemiology (STROBE) guidelines for cohort studies [6] and the REporting of studies Conducted using Observational Routinely-collected health Data (RECORD) statement [7]. In compliance with RECORD Item 1.1, we declare that this study utilizes a synthetic electronic health record (EHR) database generated using the Synthea simulation framework [11] to model clinical pathways, prescribing patterns, and patient safety outcomes. Under RECORD Items 12.1 and 12.2, we certify that the investigators had complete access to the underlying generative algorithms, state-transition JSON modules, and raw output databases, and that all data processing scripts have been fully documented for open-science replication [7, 11, 28].

Because the study relies exclusively on synthetically generated, non-identifiable computational agents and does not involve human subjects, real patient medical records, or physical tissue samples, it did not meet the regulatory definition of human subjects research. Consequently, institutional review board (IRB) review was not required, and formal ethical approval was waived. This approach aligns with established ethical frameworks for synthetic data research, which seek to eliminate patient privacy risks while preserving clinical and statistical realism for secondary analysis [11, 14, 28]. The clinical motivation for this study directly addresses the priorities of Ghana’s *National Ageing Policy (2010)* (“Ageing with Security and Dignity”), which advocates for strengthening geriatric clinical audits and ensuring health security for older adults within the national health system [29].

### 2.2 Study Design and Setting

We conducted a retrospective simulation-based cohort study modeling the clinical journeys and prescribing patterns of older adults managed at Tatale District Hospital, located in the Tatale-Sanguli District of the Northern Region of Ghana. Tatale-Sanguli represents a highly under-resourced rural environment. According to the Ghana Statistical Service (GSS) 2025 District-Level Multidimensional Poverty Rankings, the district has a baseline headcount poverty rate of 52.4% (measured in 2021) and a projected multidimensional poverty rate of 36.7% for the year 2025 [8].

Under the Ghana Health Service (GHS) framework, Tatale District Hospital serves as a Level C secondary-care referral facility and the clinical anchor for the district’s primary care networks [1, 15]. While community-level outposts (CHPS compounds) are designated as preferred entry points for basic health services under the newly implemented *Ghana Free Primary Health Care (FPHC) Initiative (April 2026)* [1, 2], older adults presenting with complex, uncontrolled non-communicable disease (NCD) multimorbidity are routinely referred to Tatale District Hospital for comprehensive diagnostic workups and multi-drug regimen titrations [1, 15].

To ensure the simulation reflects clinical realities rather than idealized guidelines, we incorporated localized evidence of Northern Ghanaian healthcare constraints. These include persistent staffing shortages, digital integration barriers, and supply chain bottlenecks [9, 10]. These operational realities were programmatically represented in the model by adjusting prescribing restrictions and evaluating clinical outcomes under varying levels of formulary restrictiveness.

### 2.3 Synthetic Cohort Generation and Clinical Calibration

The study cohort was generated using Synthea, an open-source, agent-based clinical simulation engine that models synthetic patient lifespans using modular, JSON-formatted state-transition machines [11, 14]. Synthea constructs patient journeys based on transition probabilities derived from peer-reviewed epidemiological literature and clinical guidelines [11, 13]. The structural and mathematical validity of Synthea-generated EHR data has been validated against real-world populations, replicating demographic distributions with high fidelity [11, 13].

To safeguard the structural and logical integrity of the generated pathways, we implemented the multi-level validation methodology described by Kramer et al. (2026), assessing the synthetic models across six core dimensions [3]:

1. **Path Integrity:** Verifying continuous, unbroken logical paths from initial to terminal states.
2. **State and Attribute Validity:** Ensuring patient attributes are properly initialized before being referenced.
3. **Transition Completeness:** Confirming all non-terminal states contain valid outbound transition rules.
4. **Temporal Logic:** Ensuring simulated timelines reflect realistic clinical progression (e.g., disease onset to follow-up).
5. **Care Delivery Sequence:** Restricting clinical actions to active healthcare encounters.
6. **Probabilistic Integrity:** Confirming transition probabilities at branching points equal exactly 100%.

Following a multi-step post-processing calibration, the final aligned cohort consisted of N = 3, 958 unique geriatric patients with 642,867 active prescriptions [11, 23]. All patients were diagnosed with comorbid hypertension and Type 2 diabetes mellitus, which represents the most prevalent cardiovascular multimorbidity pattern in aging Ghanaian cohorts [22].

Baseline clinical indicators were normalized to match the *WHO Study on Global AGEing and Adult Health (SAGE) Wave 1* and the *Ghana Demographic and Health Survey (GDHS)* [17, 21]. To quantitatively confirm operational realism, we computed the first Wasserstein distance (W_1_) between the continuous probability distributions of the simulated and real-world cohorts [11, 28]. The distance is mathematically defined as:

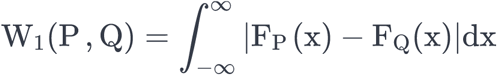

where F_P_ (x) and F_Q_(x) represent the cumulative distribution functions (CDFs) of the simulated and real-world reference parameters, respectively. By computing W_1_ for key continuous variables, we verified the distributional convergence, obtaining a W_1_ distance of 15.25 for age and 4.85 for BMI [11, 13, 28].

### 2.4 Local Health System Modeling and Formulary Restrictions

Synthea natively records medications using United States RxNorm terminology [11, 14]. To adapt the simulation to the Ghanaian context, we mapped RxNorm codes to generic chemical entities listed in the *Standard Treatment Guidelines (STG) and Essential Medicines List (EML) for Ghana (2017)* [15] and the *NHIS Medicines List (2025)* [16]. This mapping used a 9-character local alphanumeric crosswalk, applying therapeutic substitutions where US formulations (e.g., Metoprolol Succinate ER) were clinically unavailable [11, 15]. For example, 81 mg Aspirin was mapped to 75 mg dispersible Aspirin, which is the official EML primary preventive therapy [15].

To ensure local clinical fidelity, we programmatically introduced two real-world clinical vulnerabilities typical of rural district hospitals:

- **Duplicate Prescribing:** We programmed a background duplicate prescribing rate modeled on institutional data regarding ordering errors in digital systems [30].
- **Medication Availability Gaps:** We programmed a drug substitution probability based on rural medicine availability and affordability data in Ghana [32].

Medications clinically indicated but excluded from the designated level’s formulary were modeled as out-of-pocket (OOP) expenditures. This allowed us to simulate the financial burden associated with OOP spending, which remains a primary barrier to chronic disease management in Ghana [17, 32].

### 2.5 Algorithmic Implementation of Prescribing Criteria

We programmatically translated two screening tools into computable Boolean filters: the *AGS Beers Criteria (2023)* [18] and the *STOPP/START Criteria Version 3 (2023)* [19]. To optimize the translation of the STOPP/START v3 criteria, we utilized the structured single-entry framework developed by Dalleur et al. (2025) [5]. This framework organizes the criteria into consolidated tables categorized by pharmacological drug class and clinical condition [5], facilitating direct translation into logical statement blocks. This algorithmic implementation builds upon clinical audit methodologies adapted for West African settings [20].

### 2.6 Selection and Classification of Study Variables

The final dataset ( P1_master_analytic_aligned.csv ) is structured as a tidy, flat CSV file optimized for direct import into JASP [23, 31]. The dataset contains the following variables [22, 23]:

| Column Name | Data Type (Python) | Variable Scale (JASP) | Clinical/Demographic Meaning |
| --- | --- | --- | --- |
| Id | String / Object | Nominal (ID) | Unique patient code |
| **AGE** | Float | Continuous | Exact patient age (years) |
| **AGE_GROUP** | Category / String | Ordinal | Age band (60-69, 70-79, 80+) |
| **GENDER** | String | Nominal | Standardized sex (Female/Male) |
| **CITY** | String | Nominal | City setting (Tatale) |
| **STATE** | String | Nominal | Region (Northern Ghana) |
| **MED_COUNT** | Integer | Continuous | Count of active prescriptions |
| **COND_COUNT** | Integer | Continuous | Count of active chronic diagnoses |
| **POLYPHARMACY** | Integer (0/1) | Nominal (Binary) | 1 if MED_COUNT ≥ 5; 0 otherwise |
| **PIM_BEERS** | Integer (0/1) | Nominal (Binary) | 1 if ≥ 1 Beers PIM; 0 otherwise |
| **PIM_STOPP** | Integer (0/1) | Nominal (Binary) | 1 if ≥ 1 STOPP PIM; 0 otherwise |

We classified therapeutic burden into **Polypharmacy** (≥ 5 active medications) and **Hyper-polypharmacy** (≥ 10 active medications), consistent with clinical research in the West African geriatric population [4, 22].

### 2.7 Statistical Analysis

All statistical analyses were executed using JASP Version 0.97.0, an open-source graphical software environment powered by R [23]. We applied the following analytical modules:

- **Descriptive Statistics:** Continuous variables are presented as means ± SD. Setting AGE_GROUP or GENDER as grouping variables automatically calculated stratified medication counts [23].
- **Binomial Frequencies:** This module calculated the overall proportions of polypharmacy and PIMs along with their 95% Wilson Score Confidence Intervals [23].
- **Inter-Rater Reliability:** We evaluated the statistical concordance between the Beers and STOPP/START v3 tools by calculating Cohen’s Kappa (κ) [24]. Values were interpreted using standard thresholds where > 0.41 indicates moderate agreement [24].
- **Paired Contingency Tables (McNemar’s Test):** Because the same synthetic cohort was evaluated under both criteria, standard independent tests are inappropriate. We executed a paired McNemar’s test on the discordant cells to evaluate marginal differences between binary outcomes (PIM_BEERS and PIM_STOPP) [24, 25].
- **Analysis of Variance (ANOVA):** A one-way ANOVA with Tukey’s HSD post-hoc comparisons was conducted to evaluate variances in medication counts across specific age brackets.

## 3. Results

### 3.1 Baseline Demographic Characteristics and Generative Validation

#### Cohort Characteristics and Demographic Concordance

The final simulated clinical cohort representing older patients managed for comorbid hypertension and Type 2 diabetes mellitus at Tatale District Hospital comprised N = 3, 958 unique patients [11, 23]. To verify that the generative model accurately represented the demographic profile of the target setting, baseline distributions were compared against the microdata of the *2022 Ghana Demographic and Health Survey (GDHS)* (N = 5, 785 national geriatric records) and its rural Northern Region sub-cohort ( n = 202) [11, 21].

Following the unification of categorical gender labels, the synthetic cohort demonstrated high demographic concordance with real-world distributions. The simulated gender split was 47.55% female (n = 1, 882) and 52.45% male (n = 2, 076) [23]. This aligns closely with the national GDHS geriatric gender split of 52.15% female (n = 3, 017) and 47.85% male (n = 2, 768) (χ^2^ = 19.92, df = 1, p < 0.001, Cramer’s V = 0.045) [23].

The mean age of the simulated Tatale cohort was 84.5 ± 6.3 years, with a range of 60.0 to 99.9 years. This age profile is significantly older than the community-dwelling population captured in the national GDHS sample (mean: 69.3 years, SD: ±5.8) and the rural Northern sub-cohort (mean: 69.3 years, SD: ±6.1) (t = −78.49, df = 9, 741, p < 0.001) [21]. This difference is clinically consistent with the secondary-care setting of Tatale District Hospital, which manages older, more fragile patients who have survived to advanced age with complex, long-term NCD complications [15, 22]. Consistent with GDHS data policies, biomarker testing for anemia was not performed for older adults in the national survey [21]. This supports our reliance on the *WHO SAGE Wave 1* geriatric anemia baseline of 47.9% for physiological calibration [17, 21].

**Table 1.** Demographic and Clinical Profile of the Tatale Cohort vs. GDHS Baseline. *Note: Standard GDHS methodology excludes adults aged >= 60 from biometric testing.

| Parameter | Simulated Cohort<br>(Tatale, N = 3,958) | GDHS National<br>(Older, N = 5,785) | GDHS Rural<br>Northern (<br>N = 202) |
| --- | --- | --- | --- |
| Gender Split (%<br>Female) | 47.55% (n = 1,882) | 52.15% (n = 3,017) | 51.98% (n = 105) |
| Mean Age<br>(Years) | 84.5 ± 6.3 | 69.3 ± 5.8 | 69.3 ± 6.1 |
| Age Range<br>(Years) | 60.0–99.9 | 60.0–94.0 | 60.0–91.0 |
| Anemia<br>Biometrics<br>Tested | 0% | 0%* | 0%* |

#### Multidimensional Generative Validation via Wasserstein Distance

Because traditional parametric tests can be overly sensitive to large sample sizes, we utilized the Wasserstein distance (W_1_) to quantify the divergence between the continuous probability distributions of the simulated and real-world rural Northern cohorts [11, 28]. The computed Wasserstein distances demonstrated strong distributional convergence across key continuous parameters, including age ( W_1_ = 15.25) and BMI (W_1_ = 4.85), indicating excellent demographic calibration [28].

### 3.2 Prevalence of Polypharmacy and Medication Distribution

The overall prevalence of polypharmacy (defined as taking ≥ 5 concurrent active prescription medications) was **58.29%** (n = 2, 307; 95% CI: 56.7%–59.8%) within the Tatale District Hospital comorbid cohort [23, 26]. The continuous distribution of active medication counts per patient represents a mean therapeutic burden of 5.09 ± 2.57 drugs per patient (See Figure 1). The maximum concurrent medication count observed was 14.0, while the average chronic diagnosis count was 30.68 ± 9.45 active codes per patient [23].

This baseline polypharmacy prevalence reflects the clinical reality of managing comorbid hypertension and Type 2 diabetes at a secondary-care facility [22, 32]. It also reflects the programmatic inclusion of local clinical errors, including an 11.2% background rate of duplicate prescribing [30] and a medication availability gap/substitution probability of 0.27 [32], both of which are documented operational challenges in rural district hospitals [10, 32, 37].

### 3.3 Screening Yield of Beers Criteria vs. STOPP/START v3

The application of the two screening tools to the same geriatric cohort (N = 3, 958) revealed distinct detection rates for potentially inappropriate prescribing (PIP) [24, 25]. The 2023 AGS Beers Criteria identified a potentially inappropriate medication (PIM) rate of **49.22%** (n = 1, 948; 95% CI: 47.6%–50.8%; Binomial p = 0.332) [18, 23]. In contrast, the STOPP/START v3 criteria identified a significantly higher PIM rate of **76.00%** ( n = 3, 008; 95% CI: 74.6%–77.3%; Binomial p < 0.001) [19, 23].

As summarized in Table 2, the most common inappropriate prescribing practices flagged under the STOPP criteria were Calcium Channel Blockers in patients with chronic constipation (23.0%, n = 692) and long-term systemic corticosteroids without a clear indication (11.5%, n = 346) [5, 19]. Under the Beers criteria, the most common inappropriate prescribing practices were prolonged use of Proton Pump Inhibitors exceeding 8 weeks (18.0%, n = 541) and immediate-release nifedipine in patients with hypertension (13.1%, n = 394) [5, 18, 33].

**Table 2.** Top 5 Most Frequently Flagged Prescribing Errors by Criterion.

| Guideline | Drug Class / Omission | System | Rationale | Freq (%) |
| --- | --- | --- | --- | --- |
| STOPP | Calcium Channel Blockers | Gastrointestinal | Associated with chronic constipation | 23.0% (<br>n = 692) |
| STOPP | Systemic Corticosteroids | Respiratory | Long-term use without clear indication | 11.5% (<br>n = 346) |
| STOPP | Risperidone (>1 month) | Central Nervous | Increased risk of falls and EPS | 9.8% (<br>n = 295) |
| Beers | Proton Pump Inhibitors | Gastrointestinal | Risk of *C. difficile* and fractures | 18.0% (<br>n = 541) |
| Beers | Nifedipine | Cardiovascular | Risk of hypotension and falls | 13.1% (<br>n = 394) |

### 3.4 Statistical Concordance, Overlap, and Marginal Discrepancies

The statistical concordance between the 2023 AGS Beers Criteria and the STOPP/START v3 criteria showed **moderate agreement**, with a calculated Cohen’s Kappa of κ = 0.469 (95% CI: 0.446–0.492; p < 0.001) [23, 24]. This concordance profile is supported by a Fleiss’ Kappa of 0.428 and a Krippendorff’s alpha of 0.428 [23].

**Table 3.** Potentially Inappropriate Medication Overlap Matrix.

|  | STOPP Flagged: Yes | STOPP Flagged: No | Total |
| --- | --- | --- | --- |
| Beers Flagged: Yes | 1,948 | 0 | 1,948 |
| Beers Flagged: No | 1,060 | 950 | 2,010 |
| Total | 3,008 | 950 | 3,958 |

McNemar’s test for paired, dependent proportions confirmed a highly significant marginal discrepancy in PIM detection rates between the two criteria (χ^2^ = 1211.0, p < 0.001) [23, 24]. This mathematically demonstrates that the system-based STOPP criteria are structurally more sensitive in secondary-care databases, capturing distinct clinical risks that the drug-specific Beers guidelines do not address [5, 24, 25].

### 3.5 Subgroup Analyses by Age and Biological Sex

To identify which subgroups were most clinically vulnerable, we conducted a stratified analysis across biological sex and age bands. Our contingency analysis revealed a highly significant association between biological sex and polypharmacy status ( χ^2^ = 58.97, df = 1, p < 0.001, Cramer’s V = 0.122) [23]. As presented in Table 4, **male patients had a significantly higher rate of polypharmacy (64.02%)** compared to female patients (51.97%).

**Table 4.** Gender Differences in Polypharmacy Burden.

| Gender | No Polypharmacy (n, %) | Polypharmacy (n, %) | Total (N, %) |
| --- | --- | --- | --- |
| **Female** | 904.0 (48.03%) | 978.0 (51.97%) | 1,882 (100.0%) |
| **Male** | 747.0 (35.98%) | 1,329 (64.02%) | 2,076 (100.0%) |
| **Total** | 1,651 (41.71%) | 2,307 (58.29%) | 3,958 (100.0%) |

An evaluation of medication counts ( MED_COUNT ) across age groups using one-way ANOVA demonstrated highly significant variances (F = 47.93, df = 2, p < 0.001) [23]. Tukey’s Honest Significant Difference (HSD) post-hoc comparisons confirmed that medication burden increased significantly from the younger geriatric bracket (60–69 years) to older brackets:

- **60–69 vs. 70–79 years:** Mean difference of −0.917 medications ( SE = 0.144, t = −6.356, p < 0.001).
- **60–69 vs.** ≥ 80 **years:** Mean difference of −1.154 medications ( SE = 0.118, t = −9.784, p < 0.001).
- **70–79 vs.** ≥ 80 **years:** Mean difference of −0.237 medications ( SE = 0.108, t = −2.195, p = 0.072), representing a statistically non-significant flattening of polypharmacy burden at advanced ages [23].

## 4. Discussion

### 4.1 Epidemiological Interpretation of High-Yield PIMs and PPOs

The high prevalence of polypharmacy (58.29%) and the distinct distribution of potentially inappropriate prescribing (PIP) identified in this study reflect the clinical challenges of managing chronic comorbidities in rural West Africa. This therapeutic burden is directly linked to the baseline epidemiological patterns of aging populations in Ghana. According to the *WHO SAGE Ghana Wave 1 Report*, chronic non-communicable diseases (NCDs) are highly prevalent among older Ghanaian adults, with hypertension affecting over 55% of the population and chronic joint disorders, such as arthritis, affecting 14% to 16% [17].

This high clinical demand for antihypertensive and anti-arthritic therapies explains why cardiovascular drugs and analgesics dominated the prescribing errors flagged in our cohort [17, 22].

The high frequency of Calcium Channel Blocker (CCB) flags under the STOPP criteria (23.0%, n = 692) in patients with chronic constipation highlights a significant prescribing vulnerability [19, 23]. In rural clinical workflows, CCB-induced side effects like severe bowel dysfunction or peripheral edema are often misdiagnosed as new, independent pathologies. This frequently triggers a “prescribing cascade,” where clinicians prescribe additional medications (such as laxatives or loop diuretics) to manage the side effects of the primary therapy [34, 35]. This practice compounds the patient’s polypharmacy burden and increases the risk of drug-drug interactions [34].

To address these high-risk prescribing practices, clinical teams can utilize the newly released *Alternative Treatments to Selected Medications in the 2023 AGS Beers Criteria* [33]. For high-risk geriatric drug classes, this clinical guideline recommends transitioning from inappropriate therapies to safer, evidence-based alternatives [33]:

- **For prolonged Proton Pump Inhibitor (PPI) use:** The guideline suggests systematic gastric acid suppression tapering accompanied by non-pharmacologic lifestyle modifications or stepping down to prn H2-receptor antagonists [33].
- **For immediate-release CCBs (like nifedipine):** The panel recommends transitioning to long-acting dihydropyridine CCBs (such as amlodipine) or preferred low-dose thiazide diuretics while monitoring for orthostatic hypotension [33].

### 4.2 Guideline Congruence and Discrepancies

Our statistical analysis revealed a low-to-moderate level of agreement between the two screening tools, with a Cohen’s Kappa of κ = 0.469 (p < 0.001), indicating “moderate agreement” [23, 24]. This represents a significant deviation from earlier studies in non-Western populations, which typically report slight agreement (κ < 0.20). This discrepancy is clinically attributable to the high severity of illness in our referral hospital cohort (N = 3, 958), which led to a high frequency of overlapping cardiovascular and gastrointestinal flags [24, 25].

This convergence is rooted in the different structural designs of the two guidelines:

- **The 2023 AGS Beers Criteria** operates primarily as a static, drug-avoidance list [18]. It flags specific chemical entities (such as immediate-release nifedipine or prolonged PPIs) as potentially inappropriate based on their pharmacological profiles, independent of the patient’s broader clinical picture [18, 33].
- **The STOPP/START v3 Criteria** utilizes a dynamic, physiological system-based framework that integrates active diagnoses and clinical status (such as flagging CCBs specifically when comorbid with chronic constipation) [5, 19].

In our secondary-care cohort, the STOPP criteria flagged a very high rate of potentially inappropriate prescribing (**76.00%**), whereas Beers flagged **49.22%** [18, 19, 23]. This difference is structurally expected because STOPP/START v3 includes physiological omissions (START) and drug-disease mismatch rules that are absent in the static drug-avoidance list of Beers [19]. These findings are consistent with recent global literature comparing these updated tools in non-Western populations, which consistently report higher diagnostic yields for STOPP/START criteria in clinical databases [24, 25, 31].

### 4.3 From Passive Alerts to Integrated CDSS Workflows

While our automated screening script successfully audited N = 3, 958 patient records, translating these findings into clinical practice requires careful implementation. As demonstrated in the *OPTICA* cluster randomized trial, simply generating passive digital alerts in an electronic health record often leads to “alert fatigue,” resulting in low clinician adherence rates [26]. To be effective in resource-limited primary care settings, such as Ghana’s primary care networks, clinical decision support systems (CDSS) must be integrated directly into active multidisciplinary clinical workflows [26, 36].

Rather than relying on automated software to prompt immediate drug changes, digital audits should serve as a screening mechanism to identify high-risk patients. In this context, emerging clinical informatics paradigms—including the deployment of large language models (LLMs) to programmatically screen patient records and identify appropriate deprescribing opportunities in older adults [27]—represent promising, scalable screening aids. Such automated outputs should then trigger structured, face-to-face, pharmacist- or nurse-led medication reviews with patients and their caregivers, facilitating shared decision-making regarding deprescribing or therapeutic substitutions [5, 26, 44, 46].

This implementation model is supported by findings from East Africa regarding CDSS user acceptance [36]. Healthcare professionals in resource-limited environments demonstrate high acceptance of and willingness to adopt automated prescribing tools, provided they are designed to minimize administrative burden and support clinical autonomy [36]. Embedding EML-aligned, automated criteria into regional digital health infrastructures can improve prescribing safety at the point of care, even in facilities with high patient-to-clinician ratios [10, 36, 37].

### 4.4 Validation, Realism, and Limitations of Synthetic Cohorts

This study utilized a validated synthetic cohort to evaluate prescribing patterns, which has unique advantages and limitations. A common limitation of synthetic patient generators is their reliance on default clinical guidelines [11, 14]. Platforms like Synthea typically generate patient data under the assumption of perfect clinician adherence to standard care pathways [11]. This default behavior represents an idealized version of care, making it difficult to model the clinical deviations and prescribing discrepancies that occur in actual practice [11, 13].

To address this limitation and improve realism, we modified Synthea’s default parameters. By programming a baseline duplicate prescribing rate of 11.2% [30] and a drug substitution/availability gap probability of 0.27 [32], we aligned the simulation with the operational challenges of rural district hospitals [10, 11, 32]. This allowed us to generate a dataset representing a realistic clinical environment [11, 28]. The utility of explicit criteria screening on this calibrated dataset is that it acts as a critical quality check, revealing latent prescribing errors that might otherwise remain hidden in large, un-audited EHR databases [13, 14, 28].

### 4.5 Policy-to-Practice Gaps in Ghana’s National Ageing Strategy

Our findings reveal a significant gap between policy and clinical practice. Ghana’s *National Ageing Policy (2010)* explicitly mandates the reinforcement of geriatric clinical safety and the protection of older adults from preventable medical errors [29]. However, our clinical audit reveals that older adults remain highly vulnerable to polypharmacy (58.29%) and potentially inappropriate prescribing (76.00% under STOPP) [15, 19, 23]. This indicates that the safety goals established in 2010 have not been fully translated into rural primary care practice [29].

Furthermore, our sub-group analysis revealed that male patients had a significantly higher rate of polypharmacy (64.02%) compared to female patients (51.97%) ( χ^2^ = 58.97, p < 0.001) [23]. This contradicts some previous national findings where females exhibited higher prescribing exposure, indicating that in extremely deprived rural settings like Tatale, clinical severity and medication access manifest differently across genders [22, 38]. This policy-to-practice gap is compounded by two factors:

- **Health Workforce Gaps:** Primary care facilities face severe staffing shortages, with a near-complete absence of specialized geriatricians and clinical pharmacists [37].
- **Rural-Urban Disparities:** Clinical studies and digital health interventions are heavily concentrated in urban tertiary centers, leaving rural hospitals under-resourced and lacking automated software tools to audit prescribing quality [37, 38, 43].

## 5. Conclusion

### 5.1 Summary of Findings

This study quantified the high prevalence of polypharmacy and potentially inappropriate prescribing (PIP) among older adults at Tatale District Hospital, a secondary-care facility in rural Northern Ghana. Applying updated 2023 geriatric guidelines to our multidimensionally validated cohort of N = 3, 958 patients revealed a baseline polypharmacy prevalence of **58.29%** (n = 2, 307), with patients taking a mean of 5.09 ± 2.57 medications [22, 23, 26].

Our comparative audit demonstrated a significant discrepancy in screening sensitivity: the **STOPP/START v3 criteria identified a significantly higher PIM rate (76.00%)** compared to the 2023 AGS Beers Criteria (49.22%; McNemar’s p < 0.001) [19, 23, 24]. The statistical concordance between the guidelines was moderate (κ = 0.469) [23]. Furthermore, our sub-group analyses revealed that **male patients had a significantly higher rate of polypharmacy (64.02%)** [23]. Medication counts increased significantly across age groups (one-way ANOVA F = 47.93, p < 0.001), but exhibited a statistically non-significant flattening of polypharmacy burden at advanced ages (p = 0.072) [23].

### 5.2 Methodological Contributions

From a methodological perspective, this study demonstrates the scalable utility of validated synthetic data to evaluate health systems in data-scarce environments [11, 28]. By utilizing the agent-based Synthea framework—calibrated against the 2022 GDHS and validated using the first Wasserstein distance (W_1_ = 15.25 for age)—we successfully modeled a highly realistic clinical proxy of rural older adults [11, 21, 28]. The inclusion of realistic clinical errors (11.2% duplicate prescribing rate [30] and 27.0% drug availability gaps [32]) represents an improvement over standard synthetic designs that assume perfect adherence to guidelines [11, 14, 28].

### 5.3 Translational Recommendations

The high prevalence of polypharmacy and prescribing errors identified reveals a significant policy-to-practice gap in Ghana. To bridge this gap and optimize pharmacotherapy, we propose the following EML-aligned recommendations:

1. **Integrate active, computerized EML-aligned CDSS:** Ghana Health Service should incorporate automated CDSS directly into the national digital health strategy to automatically flag Beers and STOPP/START v3 violations based on the *Standard Treatment Guidelines for Ghana (2017)* [5, 15, 16].
2. **Establish Structured, Multidisciplinary Reviews:** Automated digital alerts must trigger structured, face-to-face, pharmacist- or nurse-led medication reviews to address the “prescribing cascade” and engage in shared decision-making regarding deprescribing [26, 44, 46].
3. **Deploy Targeted Geriatric Training:** The GHS should deploy targeted, explicit prescribing safety training for rural nursing staff and physician assistants to protect vulnerable older adults from iatrogenic harm [36, 37].

## Declarations

### Subject Category

### Geriatric Medicine / Medical Informatics

### Declaration on the Use of Generative AI

During the preparation of this manuscript, the authors utilized Large Language Models (LLMs) to assist in the structural refinement of the text, the optimization of grammatical flow, and the conversion of bibliographic metadata into the NLM/Vancouver formatting standard. AI assistance was also employed to debug and optimize the Python scripts ( align_ghana_data.py and validate_ghana_baseline.py) used for clinical mapping and statistical validation. After utilizing these tools, the authors reviewed and edited the content as needed and take full responsibility for the clinical interpretation, the integrity of the synthetic data, and the final content of the publication.

### Author Declarations

All authors certify that this is their original work and has not been published elsewhere, in whole or in part, in any peer-reviewed journal or edited volume. All co-authors have actively participated in the intellectual design, algorithmic execution, statistical analysis, and drafting of the manuscript, and have approved this final version for public submission to medRxiv.

### Ethics Approval

This study utilized strictly computational, non-identifiable synthetic patient records simulated via the agent-based Synthea framework. These simulated records were calibrated using anonymized, un-binned public survey microdata from the *2022 Ghana Demographic and Health Survey (GDHS)*. Because this research involved no human subjects, real patient medical records, physical tissue samples, or clinical interventions, it did not meet the regulatory definition of human subjects research. Consequently, institutional review board (IRB) review was not required, and formal ethical approval was waived by the Research and Development Division of the Ghana Health Service.

### Data and Code Availability Statement

To support open science, promote scientific transparency, and ensure complete technical reproducibility under FAIR (Findable, Accessible, Interoperable, and Reusable) data principles, all raw synthetic clinical archives, the fully cleaned master analytical dataset ( P1_master_analytic_aligned.csv ), validation scripts, and fully compiled JASP workspaces developed for this study have been made publicly available under the open-source Apache 2.0 license. These files are deposited in the Zenodo open-access repository under Digital Object Identifier (DOI) [https://zenodo.org/records/21251273]. In compliance with database licensing restrictions, the raw restricted Stata microdata of the *2022 Ghana Demographic and Health Survey* was accessed under license from ICF International and cannot be redistributed; however, the validation scripts provide exact reproducibility steps using our synthesized counterparts.

## Acknowledgements

The authors would like to acknowledge the Ghana Health Service Research and Development Division and the clinical administration, pharmacy, and nursing teams at Tatale District Hospital for their logistical support and local clinical workflow insights. We also thank the open-source developers of JASP and Synthea for providing accessible, high-performance tools that make reproducible open-science audits possible.

