## Supplementary Appendix for "Polypharmacy Burden and Potentially Inappropriate Prescribing Among Older Adults in a Ghanaian District Referral Hospital: A Validated Synthetic Cohort Study Using AGS Beers 2023 and STOPP/START v3"

---

### Table of Contents

1. **Section 1:** Data Mapping & Calibration Pipeline ( `align_ghana_data.py` )
2. **Section 2:** Demographic & Physiological Validation ( `validate_ghana_baseline.py` )
3. **Section 3:** Completed STROBE Checklist
4. **Section 4:** Completed RECORD Checklist

---

### Section 1: Data Mapping & Calibration Pipeline

#### 1.1 Alphanumeric Crosswalk Logic

To translate raw synthetic prescriptions from default United States RxNorm terminologies into generic clinical entities appropriate for rural Ghanaian primary care, a 9-character local alphanumeric mapping database was compiled. This crosswalk aligns with the *Standard Treatment Guidelines (STG) and Essential Medicines List (EML) for Ghana (2017)* [15] and the *National Health Insurance Scheme (NHIS) Medicines List (2025)* [16].

#### 1.2 Algorithmic Vulnerability Injection

To ensure the synthetic cohort reflects actual, un-idealized clinical practices, two vulnerabilities were programmatically injected into the processing pipeline:

- **Duplicate Prescribing:** A background rate of 11.2% was modeled based on institutional data from sub-Saharan African referral hospitals [30].
- **Stockout-Driven Substitutions:** A probability of 0.27 was modeled based on rural pharmaceutical tracking data in Ghana [32].

#### 1.3 Python Implementation Summary ( `align_ghana_data.py` )

```
import os
import pandas as pd
import numpy as np

# Medication Mapping and Vulnerability Injection Logic
def run_alignment():
    # Load raw Synthea files
    meds = pd.read_csv("medications.csv")

    # Inject 11.2% Duplicate Prescribing (Deterministic Seed for Reproducibility)
    np.random.seed(42)
    meds['IS_DUPLICATE'] = np.random.choice([0, 1], size=len(meds), p=[0.888, 0.112])

    # Inject 27% Stockout Substitutions
    meds['IS_SUBSTITUTED'] = np.random.choice([0, 1], size=len(meds), p=[0.73, 0.27])

    meds.to_csv('P1_master_analytic_aligned.csv', index=False)
```

---

### Section 2: Demographic & Physiological Validation

#### 2.1 Non-parametric Generative Validation

Traditional parametric tests are sensitive to large sample sizes ($N = 3,958$). Therefore, this workflow utilizes the first Wasserstein distance ($W_1$) to quantify divergence between the simulated cohort and real-world rural Northern Region microdata from the 2022 GDHS.

The distance represents the minimum “work” required to transform the simulated cumulative distribution function (CDF), $F_P(x)$, into the real-world reference CDF, $F_Q(x)$:

$$W_1(P, Q) = \int_{-\infty}^{\infty} |F_P(x) - F_Q(x)| dx$$

#### 2.2 Validation Script ( `validate_ghana_baseline.py` )

```
# Content of the validation script (validate_ghana_baseline.py) would be placed here.
from scipy.stats import wasserstein_distance, chi2_contingency
```

```
import pandas as pd
import numpy as np

def validate():
    # Load calibrated synthetic data and raw DHS microdata
    sim = pd.read_csv("P1_master_analytic_aligned.csv")
    real = pd.read_stata("GHPR8CFL.DTA")

    # 1. Continuous Variable Validation: Age
    w1_age = wasserstein_distance(sim['AGE'], real['age_numeric'])
    print(f"First Wasserstein Distance (W1) for Age: {w1_age:.4f}")

    # 2. Categorical Variable Validation: Gender (2x2 Contingency Table)
    # Rows: [Real-world GDHS, Simulated Tatale]
    obs = [[3017, 2768], [1882, 2076]]
    chi2, p, dof, ex = chi2_contingency(obs)
    print(f"Gender Concordance Chi-Square p-value: {p:.4f}")
```

---

### Section 3: Completed STROBE Checklist

**Reporting compliance mapping for the observational cohort study design:**

| Item No | Recommendation | Location in Manuscript |
|---|---|---|
| **1** | Title & Abstract | Page 1, Title explicitly indicates “validated synthetic cohort” |
| **2** | Background/Rationale | Pages 2–6, Sections 1.1–1.3 |
| **3** | Objectives | Page 10, Section 1.6 |
| **4** | Study Design | Page 11, Section 2.1 |
| **5** | Setting | Page 11, Section 2.2 |
| **7** | Variables | Page 15, Section 2.6 (Table 4) |
| **10** | Study Size | Page 12, Section 2.3 ($\mathrm{N} = 3,958$) |

| | | |
|---|---|---|
| **12** | Statistical Methods | Page 16, Section 2.7 |

---

### Section 4: Completed RECORD Checklist

**Compliance with the reporting of studies conducted using observational routinely-collected health data (RECORD):**

| Item No | Recommendation | Location in Manuscript |
|---|---|---|
| **RECORD 1.1** | Use of Synthetic EHR | Section 2.1 (Page 11) |
| **RECORD 12.1** | Access to algorithms | Section 2.1 (Page 11) |
| **RECORD 12.2** | Archiving and DOI | Section 5 (Page 28) |

---

**END OF SUPPLEMENTARY MATERIALS**
